# Identifying intergenerational risk factors for ADHD symptoms using polygenic scores in the Norwegian Mother, Father and Child Cohort

**DOI:** 10.1101/2021.02.16.21251737

**Authors:** Jean-Baptiste Pingault, Wikus Barkhuizen, Biyao Wang, Laurie J. Hannigan, Espen Moen Eilertsen, Ole A. Andreassen, Helga Ask, Martin Tesli, Ragna Bugge Askeland, George Davey Smith, Neil Davies, Ted Reichborn-Kjennerud, Eivind Ystrom, Alexandra Havdahl

**Affiliations:** Division of Psychology and Language Sciences, University College London, London, United Kingdom; MRC Social, Genetic and Developmental Psychiatry Centre, Institute of Psychiatry, King’s College, London, United Kingdom; Nic Waals Institute, Lovisenberg Diaconal Hospital, Oslo, Norway; Medical Research Council Integrative Epidemiology Unit, University of Bristol, BS8 2BN, United Kingdom; Department of Mental Disorders, Norwegian Institute of Public Health, Oslo, Norway; PROMENTA Research Center, Department of Psychology, University of Oslo, Oslo, Norway; NORMENT Centre, Institute of Clinical Medicine, University of Oslo and Division of Mental Health and Addiction, Oslo University Hospital, Oslo, Norway; K.G. Jebsen Center for Genetic Epidemiology, Department of Public Health and Nursing, NTNU, Norwegian University of Science and Technology, Norway; Population Health Sciences, Bristol Medical School, University of Bristol, Barley House, Oakfield Grove, Bristol, BS8 2BN, United Kingdom

**Author notes:** Address correspondence to Dr. Pingault; Department of Psychology, 26 Bedford Way, London, WC1H 0DS. Equal first authors (Jean-Baptiste Pingault, Wikus Barkhuizen). Equal last authors (Eivind Ystrom, Alexandra Havdahl). Financial disclosure:* The authors report no financial relationships with commercial interests.

**Keywords:** MoBa, attention-deficit/hyperactivity disorder, intergenerational transmission, genetic nurture, parenting

## Abstract

**Importance:** Knowledge of the mechanisms underlying the intergenerational transmission of risk for attention-deficit/hyperactivity disorder (ADHD) symptoms can inform psychosocial interventions.

**Objective:** To investigate whether parental genetic risk factors associate with their children’s ADHD symptoms due to genetic transmission of risk or due to parental genetic liability that influences offspring ADHD via parenting environments (genetic nurture).

**Design and participants:** This study is based on the Norwegian Mother, Father and Child Cohort Study and uses data from the Medical Birth Registry of Norway. This prospective cohort study consisted of 5,405 mother-father-offspring trios recruited between 1999 – 2008.

**Exposures:** We calculated polygenic scores for parental traits previously associated with ADHD, including psychopathology, substance use, neuroticism, educational attainment and intellectual ability.

**Main outcomes and measures:** Mothers reported on their 8-year-old children’s ADHD symptoms using the Parent/Teacher Rating Scale for Disruptive Behavior Disorders.

**Results:** Maternal polygenic scores for ADHD, autism spectrum disorder (ASD), neuroticism and smoking predicted child ADHD symptoms in bivariate analyses. After jointly modelling maternal, paternal and child polygenic scores, ADHD symptoms were predicted by children’s polygenic scores for ADHD (β = 0.10; 95% CI 0.07 to 0.14), smoking (β = 0.07; 95% CI 0.03 to 0.10) and educational attainment (β = −0.09; 95% CI −0.13 to −0.05), indicating direct genetic transmission of risk. Mothers’ polygenic scores for ASD (β = 0.05; 95% CI 0.02 to 0.08) and neuroticism (β = 0.05; 95% CI 0.01 to 0.08) predicted children’s ADHD symptoms conditional on fathers’ and children’s scores, implicating genetic nurture, or effects due to population stratification or assortative mating.

**Conclusions:** These results suggest that associations between some parental traits and offspring ADHD symptoms likely reflect a nuanced mix of direct genetic transmission (ADHD, smoking and educational attainment) and genetic nurture (ASD and neuroticism). If confirmed, these findings support previous evidence that maternal ASD or neuroticism may be possible targets for intervention to help break the chain of the intergenerational transmission of ADHD risk.

## Introduction

Understanding the mechanisms underlying intergenerational transmission of risk for attention-deficit/hyperactivity disorder (ADHD) and associated symptoms is important. Different mechanisms have different implications for intervention strategies aiming to disrupt the cycle of risk across generations.^1^ Of the possible mechanisms by which parents transmit risk to children, the role of genetics has often – somewhat surprisingly – been overlooked by epidemiologists. Here, we implement a design capitalizing on genomic data from parents and their children to systematically investigate the intergenerational transmission of risk for ADHD.^2^

Observational studies have identified many parental risk factors associated with offspring ADHD, including: parental psychopathology including ADHD itself, antisocial behaviour, depression, anxiety disorders, schizophrenia, bipolar disorder and autism spectrum disorder (ASD)^3-10^; personality traits like neuroticism;^11^ intellectual disability,^9^ education and socioeconomic position;^12-15^ psychotropic drug use during pregnancy;^16,17^ and substance use including maternal smoking, alcohol consumption and cannabis use during pregnancy.^18-22^ Like all complex behavioural traits, offspring ADHD^23,24^ and these aforementioned parental traits are partially hertiable,^25^ which leaves findings from such studies vulnerable to confounding by shared genetic factors. When genetic variants transmitted from parents to their child independently affect both the parental risk factor and the child outcome, this can lead to associations between parental risk factors and ADHD symptoms in the absence of environmental pathways of causation or inflate associations to make environmental pathways appear more consequential than they are.^26^ Addressing this issue is fundamental to assess whether modifiable parental risk factors are likely to be effective targets for intervention to reduce the risk or severity of ADHD symptoms in children.

The need to account for genetic transmission in intergenerational settings has long been recognised and several genetically informed designs for causal inference^26,27^ have aimed to address it, including the adoption design,^28^ the (extended) children-of-twins design^10,15,29^ and the In Vitro Fertilization (IVF) design.^30^ Adoption and IVF designs account for intergenerational genetic transmission by comparing genetically and non-genetically related parent-child pairs and the (extended) children-of-twins design by directly estimating the role of genetics in the intergenerational transmission of risk. Findings from these genetically informed studies have clearly demonstrated that genetic transmission from parents to their children need to be accounted for in aetiological studies of childhood ADHD. However, by their very nature, such genetically informed designs require specific populations, leading to both few samples and small sample sizes when available. This also hampers generalizability of findings. Furthermore, most studies focus on one or a few putative parental risk factors at a time. Each design also has its own limitations. For example, in the IVF design, the association between ADHD symptoms in the biological mother and child ADHD symptoms might be partly due to *in utero* influences rather than genetic transmission; conversely, in the adoption design, the association between ADHD symptoms in the adoptive mother and child ADHD symptoms may be partly due to reverse causation rather than the impact of maternal ADHD symptoms. As such, triangulation of findings between studies that employ genetically informed designs with different underlying assumptions and limitations is important.

Intergenerational transmission can also be investigated in population-based samples with genotype data on parents and offspring and phenotypic measures on offspring outcomes. Instead of phenotypic parental measures, genetic variants associated with the parental risk factor (e.g. parental ADHD) are combined into a polygenic score and used in the analyses. The relative importance of different transmission mechanisms can be investigated either by splitting the parental polygenic score between variants transmitted and not transmitted to the child, ^31,32^ or by employing a statistical control design where parental and offspring polygenic scores are jointly modelled to predict offspring outcome (Figure 1). Because the nontransmitted parental alleles cannot be biologically expressed in the offspring, these designs can be used to distinguish genetic versus environmental routes of risk transmission across generations. This is analogous to what happens in non-genetically related pairs in the IVF design and in an adoption design. Associations between nontransmitted genetic influences and offspring phenotypes cannot be explained by genetic transmission and happens via an environmental route, otherwise known as genetic nurture.^32^ As an example, ADHD variants might influence parental ADHD liability, which in turn influences the child rearing environment and ultimately the child ADHD symptoms. Estimates of genetic nurture may also capture population-level effects if parental traits are under the influence of assortative mating or population stratification.

**Figure 1.**
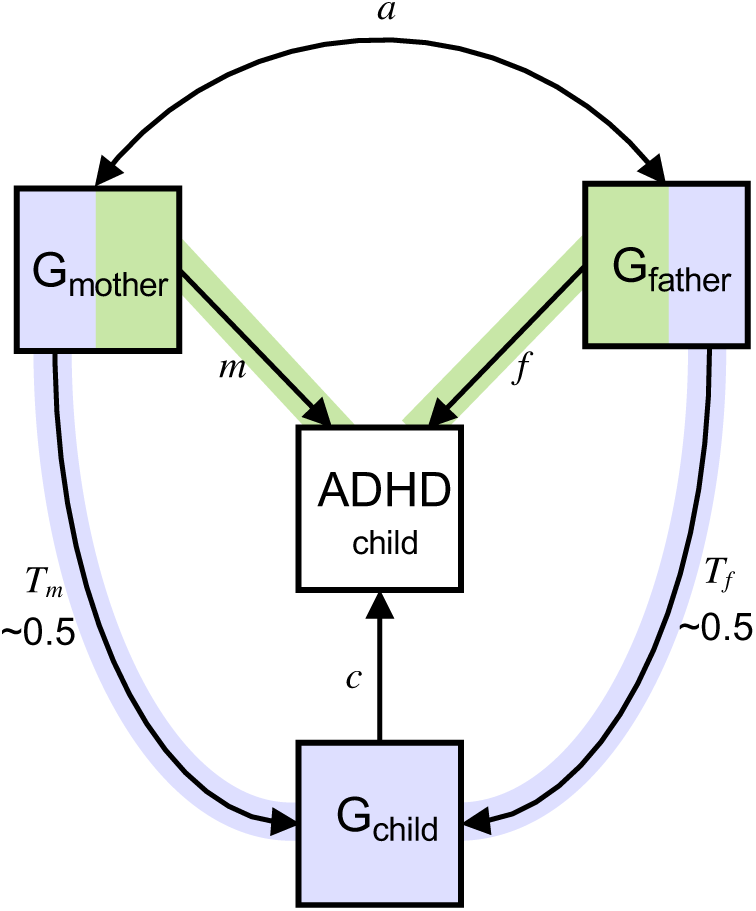
Statistical control model to investigate the intergenerational transmission of risk. *Note:* Path diagram illustrating a regression model with polygenic scores for mother (G_mother_), father (G_father_) and child (G_child_) predicting child ADHD symptoms. Genetic influences shared between the child and their mother (path *T*_*m*_) and father (path *T*_*f*_) are illustrated in purple. Coefficients for path *c* represent direct genetic transmission of risk and reflect the genetic influences of the child’s polygenic score on their ADHD symptoms after controlling for the influences of parental polygenic scores. Genetic nurture effects (highlighted in green) on childhood ADHD are estimated from path coefficients *m* for mothers and *f* for fathers. Coefficients for paths *m* and *f* reflects the effects of parental genotypes on childhood ADHD symptoms after statistically controlling for the genetic influences transmitted from parents to their child and for covariation between parental polygenic scores (path *a*). Note that path *a* will be near zero in the absence of assortative mating.

To date, only one study has used transmitted and nontransmitted polygenic scores to investigate the association between parental risk factors (ADHD and education) on child ADHD symptoms.^33^ They found that the association between these two parental traits and child ADHD symptoms could mainly be attributed to genetic transmission rather than to effects mediated by the child rearing environment. However, the study focused on just two potential risk factors and relied on a relatively small sample of children and their parents (*N* = 2518; effective sample size = 1702).

In the present study, we use a large population-based cohort of genotyped trios to systematically investigate a range of parental risk factors including psychopathology, substance use, neuroticism, educational attainment and intellectual ability. We jointly model maternal, paternal and child polygenic scores to assess the role of genetic transmission of risk from parents to offspring versus environmental routes of transmission.

## Method

### Participants and genotype data

The Norwegian Mother, Father and Child Cohort Study (MoBa)^34,35^ is a population-based pregnancy cohort study conducted by the Norwegian Institute of Public Health. Participants were recruited from all over Norway from 1999-2008. The women consented to participation in 41% of the pregnancies. The cohort now includes 114,500 children, 95,200 mothers and 75,200 fathers. Blood samples were obtained from both parents during pregnancy and from mothers and children (umbilical cord) at birth. The Medical Birth Registry (MBRN) is a national health registry containing information about all births in Norway, and was used to obtain children’s year of birth and sex. The current study is based on version 12 of the quality-assured data files released for research on January 2019. The establishment of MoBa and initial data collection was based on a license from the Norwegian Data protection agency and approval from The Regional Committees for Medical and Health Research Ethics (REK). The MoBa cohort is based on regulations based on the Norwegian Health Registry Act. The current study was approved by REK (2016/1702).

The eligible sample came from a subset of MoBa families with genotypic data^36^ available for mother-father-offspring trios. The quality control of MoBa genetic data has been described elsewhere^37^ and in the Supplementary Note. We randomly excluded one individual within pairs of relatives within and across generations and across genotyping batches, whilst prioritising those with complete genetic data within family trios. Of the resulting 11,262 families, child ADHD symptom scores at 8 years were available for 5,405 families (Supplementary Table 1).

### Measures

#### Childhood symptoms of Attention-Deficit Hyperactivity Disorder

Childhood symptoms of ADHD was reported by mothers using the Parent/Teacher Rating Scale for Disruptive Behavior Disorders (RS-DBD).^38^ The RS-DBD contains 18 items related to ADHD based on DSM-IV criteria. Items were rated on a four-point scale and coded as continuous summary scores. Participants with fewer than half completed items from the RS-DBD were excluded. ADHD symptom scores were standardized to have a mean of zero and a standard deviation of 1.

### Parental risk factors

Polygenic scores were used as measures genetic liabilities for parental risk factors. Polygenic scores are individual-level scores that summarize genetic risk for a given phenotype. The scores are calculated as the weighted sum of effect alleles for all single nucleotide polymorphisms (SNP) in an individual. Weights are obtained for each SNP in summary statistics from Genome-Wide Associations Studies (GWAS).

We derived polygenic scores for 12 parental risk factors^39-50^ (Table 1) capturing the following domains: parental psychopathology, educational attainment, intelligence and substance use. GWAS summary statistics for risk factors were selected if of sufficient power to conduct polygenic score analyses, if GWAS were conducted on samples of European decent and did not include overlapping participants with the target sample. Power was determined based on established criteria^50,51^ as follows: risk factors were excluded from the current analyses if SNP-heritability (SNP-h^2^) estimates were lower than 0.05 or if SNP-h^2^ Z-scores (calculated as SNP-h^2^ divided by its standard error) were less than 2 (as was the case for GWAS on antisocial behaviour^3^).

**Table 1.** Genome-wide association study summary statistics used to compute polygenic scores

| Phenotype | Study | *N* | Sample prevalence | Population prevalence | SNP-<br>$h^2$ | *SE* |
| --- | --- | --- | --- | --- | --- | --- |
| ADHD | Demontis *et al.* (2019) | 53,293 | 0.358 | 0.050 | 0.2167 | 0.0140 |
| ASD | Grove *et al.* (2019) | 46,351 | 0.397 | 0.005 | 0.0960 | 0.0083 |
| Schizophrenia | Pardinas *et al.* (2018) | 105,318 | 0.386 | 0.010 | 0.2342 | 0.0083 |
| Bipolar disorder | Stahl *et al.* (2019) | 51,710 | 0.396 | 0.020 | 0.2372 | 0.0119 |
| Depression | Howard *et al.* (2019) | 500,199 | 0.341 | 0.341 | 0.0999 | 0.0039 |
| Anxiety disorder | Purves *et al.* (2019) | 83,566 | 0.305 | 0.209 | 0.1645 | 0.0108 |
| Neuroticism | Nagel *et al.* (2018)<sup>a</sup> | 390,278 | - | - | 0.1030 | 0.0034 |
| Intelligence | Savage *et al.* (2018) | 269,867 | - | - | 0.1846 | 0.0077 |
| EA | Watanabe *et al.* (2019) | 318,526 | - | - | 0.1108 | 0.0044 |
| Alcohol use | Linner *et al.* (2019) | 414,343 | - | - | 0.0692 | 0.0029 |
| Smoking | Wootton *et al.* (2019) | 462,690 | - | - | 0.0943 | 0.0032 |
| Cannabis use | Pasman *et al.* (2018)<sup>a</sup> | 162,082 | 0.268 | 0.268 | 0.1199 | 0.0077 |

*Note:* SNP- $h^2$ = Single nucleotide polymorphism (SNP) heritability; ADHD = Attention-deficit/hyperactivity disorder; ASD = Autism spectrum disorder; EA = Educational attainment; Alcohol use was defined as the average number of alcoholic drinks participants drank per week; Cannabis use was assessed as a binary measure of lifetime use; Smoking was based on a lifetime smoking index and captured the heaviness and duration of tobacco smoking and included non-smokers; Additional details on these measures and method used to calculate SNP- $h^2$ are provided in the supplementary note.
<sup>a</sup> excluding 23andMe participants.

Quality control of publicly available summary statistics was performed prior to conducting polygenic score analysis following recommended procedures^51^ to exclude sex chromosomes, palindromic variants and variants with INFO < 0.8 and MAF < 0.01. Polygenic scores were computed using PRSice software,^52^ with prior clumping to remove SNPs in linkage disequilibrium (r^2^ > 0.10 within a 500kb window). Polygenic scores corresponding to each risk factor were generated in mothers, fathers and children based on all variants (*p*-value threshold of 1). Correlations between parental polygenic scores are provided in Supplementary Table 2. Supplementary Table 3 provides a comparison of mean polygenic scores in family members with and without child phenotypic measures available.

### Data Analysis

Prior to analyses, all polygenic scores were standardized to a mean of zero and a variance of one and residualized for batch effects and the 10 first principal components to control for population stratification. All models detailed below additionally controlled for child sex and year of birth. False Discovery Rate (FDR) of 5% was applied to control for multiple testing, the number of tests being equal to the number of considered risk factors multiplied by three family members. Variance inflation factors (VIF) were computed to identify the presence of multicollinearity between predictors in multivariable models, with VIF > 4 indicating multicollinearity.

#### Bivariate models

Linear regressions with child ADHD symptoms as the outcome and each polygenic score as the predictor were conducted.

#### Multivariable models: within risk factor

The within risk factor multivariable analysis jointly considered the polygenic scores for mother, father, and child (Figure 1).

#### Multivariable model: across risk factors

We then considered polygenic scores for mother, father and child for risk factors for which at least one family member’s polygenic score significantly predicted child ADHD symptoms in the within risk factor analysis, jointly in one model. We jointly modelled these risk factors to assess whether pleiotropy between these traits could affect estimates of genetic nurture and direct genetic effects.

## Results

Table 2 shows the results from the bivariate and within risk factor multivariable analyses. ADHD and smoking polygenic scores of both mother and child all predicted child ADHD symptoms in bivariate analyses, whereas in the multivariable models, only child polygenic scores remained significant predictors - suggesting direct genetic effects. The offspring ASD, neuroticism and intelligence polygenic scores were associated with ADHD symptoms in the bivariate, but not the multivariable models, providing little evidence for direct genetic effects. The maternal ASD and neuroticism polygenic scores were associated with offspring ADHD symptoms in both the bivariate and multivariable models. That is, even accounting for expression of these genetic influences in children, the maternal scores for ASD and neuroticism still significantly predicted their children’s ADHD symptoms. These associations are therefore not attributable to genetic transmission and may instead be due to genetic nurture effects of mothers’ phenotypes or other population or familial level effects. The offspring educational attainment polygenic score negatively associated with ADHD symptoms in both bivariate and multivariable models, indicating direct genetic effects. VIF between polygenic scores of family members in the multivariable models ranged from 1.44 to 2.29, indicating that multicollinearity did not affect the results.

**Table 2.**
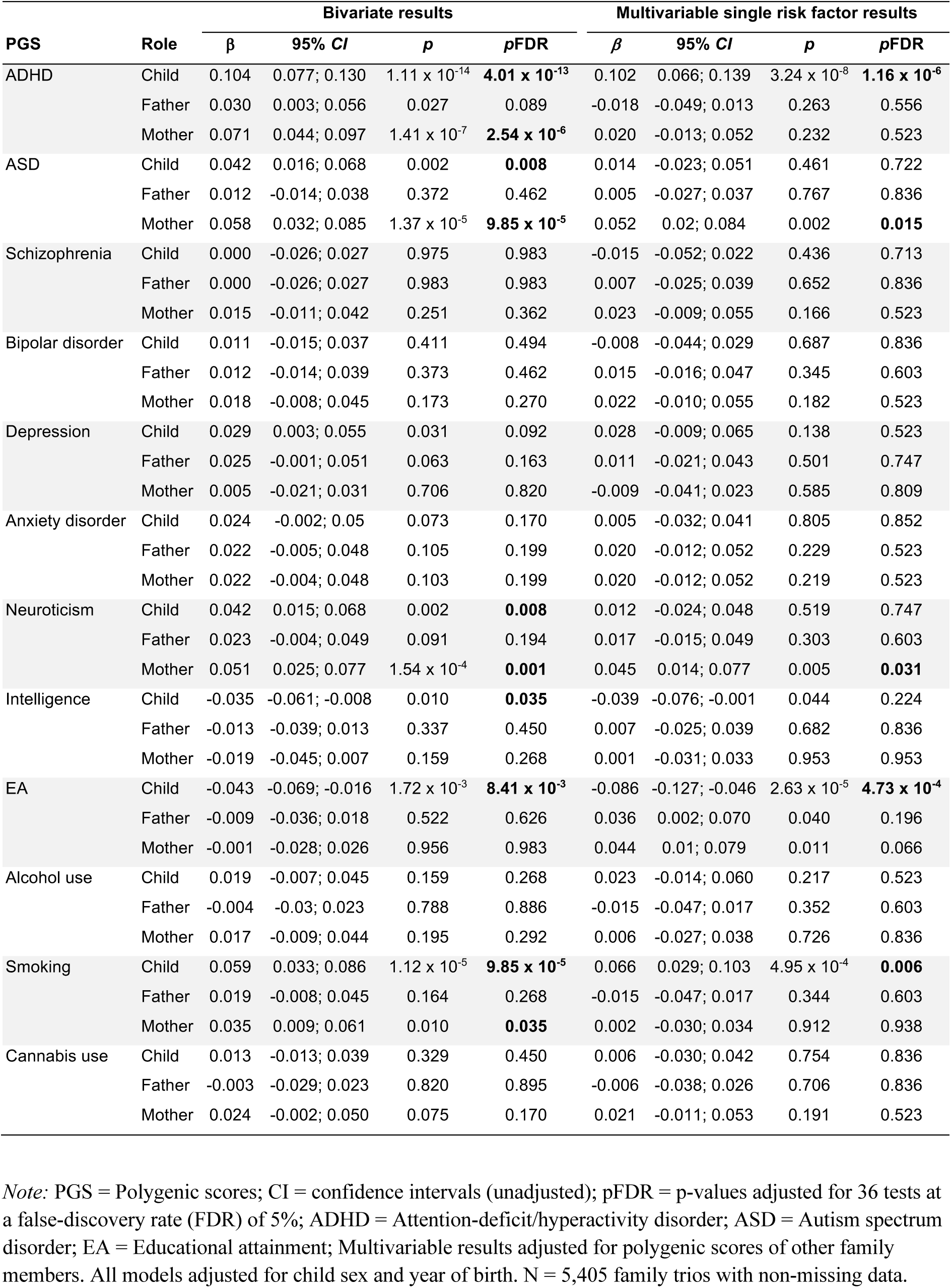
Family trios’ polygenic scores predicting child ADHD symptoms before and after adjusting for polygenic scores of other family members (within risk factors).

| PGS | Role | Bivariate results |  |  |  | Multivariable single risk factor results |  |  |  |
| --- | --- | --- | --- | --- | --- | --- | --- | --- | --- |
| | | $\beta$ | 95% CI | $p$ | pFDR | $\beta$ | 95% CI | $p$ | pFDR |
| ADHD | Child | 0.104 | 0.077; 0.130 | $1.11 \times 10^{-14}$ | **$4.01 \times 10^{-13}$** | 0.102 | 0.066; 0.139 | $3.24 \times 10^{-8}$ | **$1.16 \times 10^{-6}$** |
|  | Father | 0.030 | 0.003; 0.056 | 0.027 | 0.089 | -0.018 | -0.049; 0.013 | 0.263 | 0.556 |
| | Mother | 0.071 | 0.044; 0.097 | $1.41 \times 10^{-7}$ | **$2.54 \times 10^{-6}$** | 0.020 | -0.013; 0.052 | 0.232 | 0.523 |
| ASD | Child | 0.042 | 0.016; 0.068 | 0.002 | **0.008** | 0.014 | -0.023; 0.051 | 0.461 | 0.722 |
|  | Father | 0.012 | -0.014; 0.038 | 0.372 | 0.462 | 0.005 | -0.027; 0.037 | 0.767 | 0.836 |
| | Mother | 0.058 | 0.032; 0.085 | $1.37 \times 10^{-5}$ | **$9.85 \times 10^{-5}$** | 0.052 | 0.02; 0.084 | 0.002 | **0.015** |
| Schizophrenia | Child | 0.000 | -0.026; 0.027 | 0.975 | 0.983 | -0.015 | -0.052; 0.022 | 0.436 | 0.713 |
|  | Father | 0.000 | -0.026; 0.027 | 0.983 | 0.983 | 0.007 | -0.025; 0.039 | 0.652 | 0.836 |
|  | Mother | 0.015 | -0.011; 0.042 | 0.251 | 0.362 | 0.023 | -0.009; 0.055 | 0.166 | 0.523 |
| Bipolar disorder | Child | 0.011 | -0.015; 0.037 | 0.411 | 0.494 | -0.008 | -0.044; 0.029 | 0.687 | 0.836 |
|  | Father | 0.012 | -0.014; 0.039 | 0.373 | 0.462 | 0.015 | -0.016; 0.047 | 0.345 | 0.603 |
|  | Mother | 0.018 | -0.008; 0.045 | 0.173 | 0.270 | 0.022 | -0.010; 0.055 | 0.182 | 0.523 |
| Depression | Child | 0.029 | 0.003; 0.055 | 0.031 | 0.092 | 0.028 | -0.009; 0.065 | 0.138 | 0.523 |
|  | Father | 0.025 | -0.001; 0.051 | 0.063 | 0.163 | 0.011 | -0.021; 0.043 | 0.501 | 0.747 |
|  | Mother | 0.005 | -0.021; 0.031 | 0.706 | 0.820 | -0.009 | -0.041; 0.023 | 0.585 | 0.809 |
| Anxiety disorder | Child | 0.024 | -0.002; 0.05 | 0.073 | 0.170 | 0.005 | -0.032; 0.041 | 0.805 | 0.852 |
|  | Father | 0.022 | -0.005; 0.048 | 0.105 | 0.199 | 0.020 | -0.012; 0.052 | 0.229 | 0.523 |
|  | Mother | 0.022 | -0.004; 0.048 | 0.103 | 0.199 | 0.020 | -0.012; 0.052 | 0.219 | 0.523 |
| Neuroticism | Child | 0.042 | 0.015; 0.068 | 0.002 | **0.008** | 0.012 | -0.024; 0.048 | 0.519 | 0.747 |
|  | Father | 0.023 | -0.004; 0.049 | 0.091 | 0.194 | 0.017 | -0.015; 0.049 | 0.303 | 0.603 |
| | Mother | 0.051 | 0.025; 0.077 | $1.54 \times 10^{-4}$ | **0.001** | 0.045 | 0.014; 0.077 | 0.005 | **0.031** |
| Intelligence | Child | -0.035 | -0.061; -0.008 | 0.010 | **0.035** | -0.039 | -0.076; -0.001 | 0.044 | 0.224 |
|  | Father | -0.013 | -0.039; 0.013 | 0.337 | 0.450 | 0.007 | -0.025; 0.039 | 0.682 | 0.836 |
|  | Mother | -0.019 | -0.045; 0.007 | 0.159 | 0.268 | 0.001 | -0.031; 0.033 | 0.953 | 0.953 |
| EA | Child | -0.043 | -0.069; -0.016 | $1.72 \times 10^{-3}$ | **$8.41 \times 10^{-3}$** | -0.086 | -0.127; -0.046 | $2.63 \times 10^{-5}$ | **$4.73 \times 10^{-4}$** |
|  | Father | -0.009 | -0.036; 0.018 | 0.522 | 0.626 | 0.036 | 0.002; 0.070 | 0.040 | 0.196 |
|  | Mother | -0.001 | -0.028; 0.026 | 0.956 | 0.983 | 0.044 | 0.01; 0.079 | 0.011 | 0.066 |
| Alcohol use | Child | 0.019 | -0.007; 0.045 | 0.159 | 0.268 | 0.023 | -0.014; 0.060 | 0.217 | 0.523 |
|  | Father | -0.004 | -0.03; 0.023 | 0.788 | 0.886 | -0.015 | -0.047; 0.017 | 0.352 | 0.603 |
|  | Mother | 0.017 | -0.009; 0.044 | 0.195 | 0.292 | 0.006 | -0.027; 0.038 | 0.726 | 0.836 |
| Smoking | Child | 0.059 | 0.033; 0.086 | $1.12 \times 10^{-5}$ | **$9.85 \times 10^{-5}$** | 0.066 | 0.029; 0.103 | $4.95 \times 10^{-4}$ | **0.006** |
|  | Father | 0.019 | -0.008; 0.045 | 0.164 | 0.268 | -0.015 | -0.047; 0.017 | 0.344 | 0.603 |
|  | Mother | 0.035 | 0.009; 0.061 | 0.010 | **0.035** | 0.002 | -0.030; 0.034 | 0.912 | 0.938 |
| Cannabis use | Child | 0.013 | -0.013; 0.039 | 0.329 | 0.450 | 0.006 | -0.030; 0.042 | 0.754 | 0.836 |
|  | Father | -0.003 | -0.029; 0.023 | 0.820 | 0.895 | -0.006 | -0.038; 0.026 | 0.706 | 0.836 |
|  | Mother | 0.024 | -0.002; 0.050 | 0.075 | 0.170 | 0.021 | -0.011; 0.053 | 0.191 | 0.523 |

*Note:* PGS = Polygenic scores; CI = confidence intervals (unadjusted); pFDR = p-values adjusted for 36 tests at a false-discovery rate (FDR) of 5%; ADHD = Attention-deficit/hyperactivity disorder; ASD = Autism spectrum disorder; EA = Educational attainment; Multivariable results adjusted for polygenic scores of other family members. All models adjusted for child sex and year of birth. N = 5,405 family trios with non-missing data.

Results for the multivariable model across risk factors are provided in Table 3 and include polygenic scores for ADHD, ASD, educational attainment, neuroticism and smoking. The findings were broadly consistent with those from the within risk factor models described above and there was evidence of direct genetic effects of ADHD and educational attainment (but not smoking) on childhood ADHD symptoms as well as for genetic nurture effects of mothers’ ASD and neuroticism. No multicollinearity between polygenic score predictors was detected (VIF range 1.52 to 2.36).

**Table 3.**
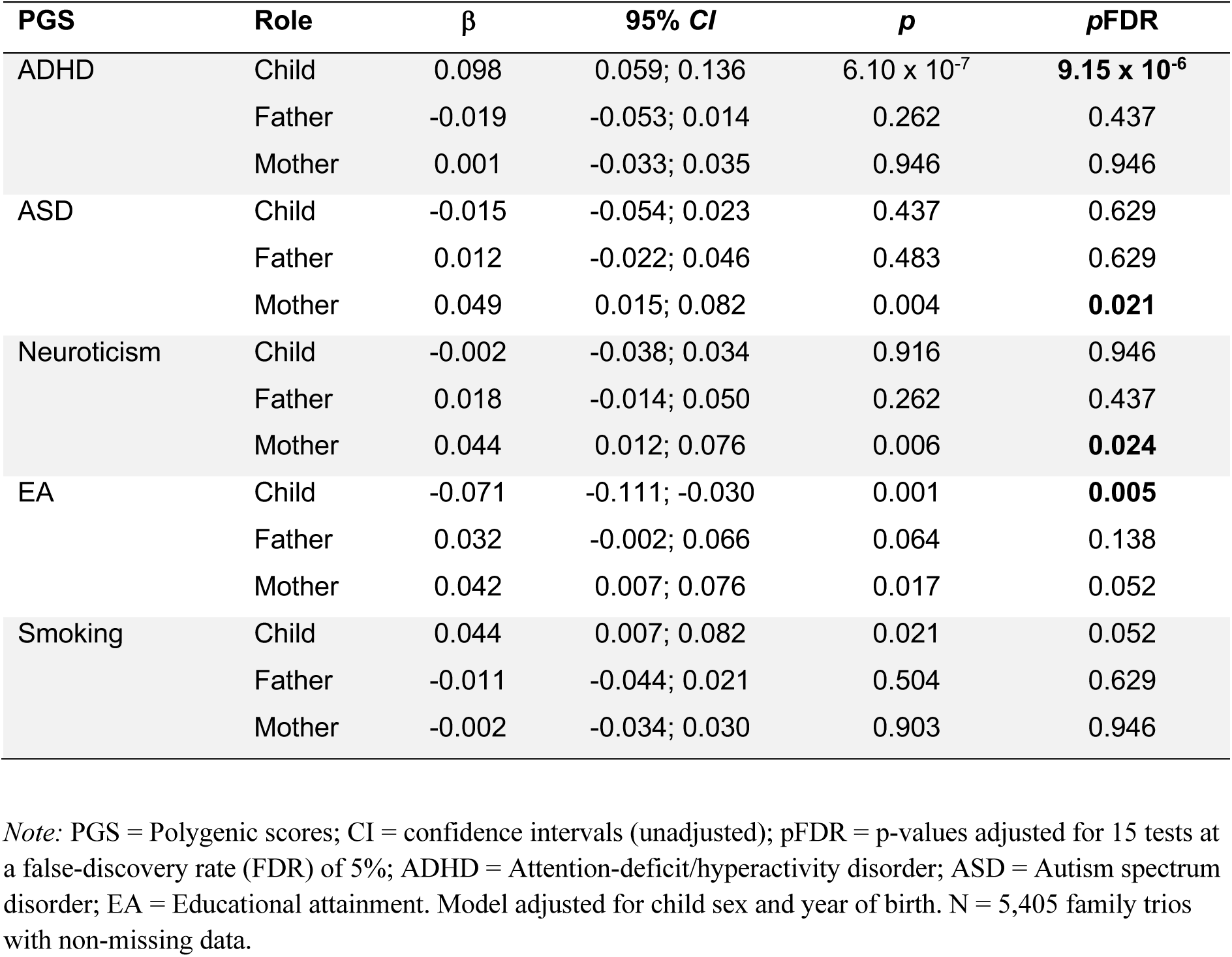
Polygenic scores predicting child ADHD symptoms adjusted for polygenic scores across multiple risk factors.

## Discussion

We examined the associations of polygenic scores for behavioural and mental health-related parental risk factors - such as psychopathology, education and substance use - and child ADHD symptoms. Specifically, we used polygenic scores for family trios to test whether associations between these parental risk factors and child ADHD symptoms could indicate genetic nurture or genetic transmission. Several parental polygenic scores associated with child ADHD symptoms after multiple testing correction, including maternal polygenic scores for ADHD itself, ASD, neuroticism and smoking. In multivariable models which included polygenic scores for offspring, mother and father, we found evidence that – of the polygenic scores indexing parental genetic liability for common risk factors – only maternal genetic predisposition to ASD and neuroticism associated with child ADHD symptoms.

### Genetic transmission versus genetic nurture

We found that maternal polygenic scores for smoking were associated with their child’s ADHD symptoms, but this association disappeared after accounting for genetic transmission. This suggests that associations between exposure to maternal smoking and child ADHD symptoms may arise, at least in part, due to genetic influences shared between mothers and their children. These findings are consistent with a large body of evidence using multiple genetically informed and causal inference methods, showing that the association between maternal smoking and child ADHD symptoms is likely due to intergenerational genetic transmission.^30,53-55^ Furthermore, our findings show that the child polygenic score for smoking is associated with ADHD symptoms in 8-year old children, i.e. prior to smoking initiation, and independently of parental genetic risk for smoking. This indicates direct genetic effects whereby genetic influences on smoking, inherited by the child, independently associate with their ADHD symptoms. Direct genetic effects could inflate or generate a spurious association between parental smoking and child ADHD symptoms. We found that the direct genetic effect of smoking was attenuated after adjusting for the polygenic scores of ADHD, ASD, neuroticism and educational attainment. Shared genetic variation may influence both smoking and these risk factors, which could explain the direct genetic association we observed between children’s polygenic liability to smoking and their ADHD symptoms.

We also found that the maternal ADHD polygenic scores associated with the child ADHD symptoms, but these associations were attenuated after adjusting for polygenic scores of other family members. This suggests that the bivariate association of parental polygenic risk for ADHD is unlikely to be mediated via the parents’ phenotype (i.e. the environment). Thus, we found little evidence that parental liability to ADHD affect their offspring’s ADHD symptoms. Rather, parental genetic risk for ADHD is directly genetically transmitted to and expressed in the child. This finding is in line with a previous study in a Dutch sample.^33^ However, this finding contrasts with an adoption study that found evidence for associations between child ADHD symptoms and ADHD symptoms in both their biological and adoptive mothers, which suggested the presence of intergenerational genetic transmission but also a possible environmentally mediated impact of maternal ADHD on child ADHD.^28^ However, the observed association between ADHD symptoms in adoptive mothers and ADHD symptoms in the child may be due to reverse causation which cannot happen in the design we used (i.e. child ADHD symptoms cannot change parental genotypes).

Our findings do not exclude the possibility that other parental characteristics affect their offspring’s development of ADHD symptoms. Indeed, we found some evidence that maternal genetic liability to ASD and neuroticism not transmitted to their offspring associated with offspring ADHD symptoms. Both autistic traits and neuroticism reported by parents have been phenotypically associated with parenting difficulties,^56,57^ which may explain the associations we found between maternal genetic variation and offspring ADHD symptoms in multivariable models. These findings suggest a potential role for parenting interventions for parents at high risk of ASD and neuroticism. However, these findings warrant replication. We found little evidence that genetic liability to other parental characteristics such as psychiatric disorders, alcohol use and intelligence were associated with offspring ADHD symptoms despite previous evidence for epidemiological associations.^3-10,18-22^ Our ability to detect associations between parental traits and offspring ADHD symptoms was limited by the power of current polygenic scores used as proxies for these parental traits.

### Implications for intergenerational psychiatry

Our findings suggest that genetic transmission may play an important role in explaining intergenerational transmission of ADHD symptoms. Genetic transmission means that phenotypes arise separately in mother and child due to genetic influences they share, but independently of the environment created by parents. Conversely, genetic nurture effects imply that some parental risk factors matter as they foster nurturing (or negative) environments that affect the child. Implications for interventions are diametrically opposed. In the case of genetic nurture, positive effects of interventions targeting parents are expected to cascade across generations and improve child outcomes. In the case of genetic transmission, benefits of interventions targeting parents are unlikely to benefit the child. Instead, direct interventions aiming to prevent ADHD symptoms in children should be prioritized. Such findings for ADHD symptoms contrast with findings for educational attainment in children where substantial genetic nurture effects have been found.^31,32^ As such, we should not be expecting universal patterns when it comes to explaining the role of intergenerational risk factors in children’s developmental outcomes. Emerging genetically informed methods^26,31,58^ should shortly render a detailed depiction of the intergenerational transmission of risk for psychiatric symptoms.

### Limitations

As in all longitudinal studies, attrition can bias findings. Results from a Norwegian cohort may not generalize to all contexts. Despite relying on the largest genotyped cohort of trios so far, power may be an issue to detect small intergenerational effects. Replications of findings using multiple imputation methods and in multiple cohorts across countries will be essential in addressing limitations such as selection bias, generalizability and power. Using polygenic scores does not enable an appropriate estimation of causal effects of intergenerational risk factors. As such, genetic nurture effects are just suggestive of plausible causal effects of the risk factors under scrutiny. Explicit causal inference methods such as Mendelian randomization (MR) should be implemented in future analyses.^18^ Although estimates from polygenic scores can be mathematically similar to MR estimates, MR offers a suite of methods more robust to confounding due to biological pleiotropy than polygenic scores. The development of MR in an intergenerational context^44-46^ will enable explicit estimations of causal effects when appropriately large samples and suitable summary statistics become available. We used a parent-rated scale of ADHD symptoms rather than a binary classification of clinically diagnosed ADHD as the outcome. Related to this, the use of genetic proxies, whether with polygenic scores or MR, captures the genetic liability for a disorder rather than the disorder itself. Although evidence consistently points towards ADHD being the extreme tail of a continuous distribution of ADHD symptoms,^47^ such methods cannot easily capture nonlinear effects (e.g. parental effects that arise when a parent is diagnosed with ADHD but are absent at sub-threshold levels). Finally, we did not consider all possible parental risk factors for ADHD here. Future projects will follow up on the intergenerational transmission of risk for child ADHD due to cardiometabolic, nutritional and immune-related parental traits.

### Conclusions

Our findings illustrate the importance of accounting for genetic influences when investigating the intergenerational transmission of ADHD risk. Genetic influences on ADHD, EA and smoking, transmitted from parents and expressed in their offspring, may confound or result in spurious epidemiological associations between these parental risk factors and offspring ADHD symptoms. Our findings suggest that genetic nurture, assortative mating or population structure may partly explain previously reported associations between maternal ASD and neuroticism and ADHD symptoms in children.

## Supporting information

STROBE

## Data Availability

Access to MoBa data requires approval from The Norwegian Institute of Public Health (NIPH).
https://www.fhi.no/en/more/access-to-data/applying-for-access-to-data/

## Acknowledgements

The Norwegian Mother, Father and Child Cohort Study is supported by the Norwegian Ministry of Health and Care Services and the Ministry of Education and Research. We are grateful to all the participating families in Norway who take part in this on-going cohort study. We thank the Norwegian Institute of Public Health (NIPH) for generating high-quality genomic data, as part of the HARVEST collaboration which is supported by the Research Council of Norway (#229624). We further thank the Center for Diabetes Research, the University of Bergen for providing genotype data and performing quality control and imputation of the data funded by the ERC AdG project SELECTionPREDISPOSED, Stiftelsen Kristian Gerhard Jebsen, Trond Mohn Foundation, the Research Council of Norway, the Novo Nordisk Foundation, the University of Bergen, and the Western Norway health Authorities (Helse Vest). This work was performed on the TSD (Tjeneste for Sensitive Data) facilities, owned by the University of Oslo, operated and developed by the TSD service group at the University of Oslo, IT-Department (USIT).

