## Supplementary for "Identifying intergenerational risk factors for ADHD symptoms using polygenic scores in the Norwegian Mother, Father and Child Cohort"

### Supplementary Note

#### *Summary statistics used to conduct polygenic scores*

*ADHD.* Summary statistics were obtained from a genome-wide association study (GWAS) on ADHD[1] from https://www.med.unc.edu/pgc/download-results/. The ADHD GWAS was performed on 19,099 ADHD cases and 34,194 controls consisting of European samples from the Lundbeck Foundation Initiative for Integrative Psychiatric Research (iPSYCH) and the Psychiatric Genomics Consortium (PGC).[1] Cases in iPSYCH were identified based on ICD-10 criteria obtained from psychiatric diagnoses obtained from a national research register. Case definitions for PCG cohorts has been described elsewhere.[2]

*Autism Spectrum Disorder (ASD).* Summary statistics were downloaded from https://ipsych.dk/en/research/downloads/ and was based on a GWAS meta-analysis of iPSYCH and PGC samples (18,382 cases and 27,969 controls).[3] Cases were based on validated registry-based diagnoses of ASD according to ICD-10 criteria and for the PGC described elsewhere.[4]

*Schizophrenia.* The GWAS summary statistics is available at http://walters.psycm.cf.ac.uk/ and was based on a meta-analysis of PGC and CLOZUK samples resulting in 40,675 schizophrenia cases and 64,643 controls.[5] Cases were identified based on DSM-IV criteria for schizophrenia or schizoaffective disorder.

*Bipolar disorder.* Summary statistics, available from the PGC (https://www.med.unc.edu/pgc/results-and-downloads) was based on a GWAS meta-analysis on 20,352 cases and 31,358 controls. Different clinical interview formats were used to diagnose bipolar disorder, described in full elsewhere.[6]

*Depression*. Summary statistics for depression was from a meta-GWAS[7] on 170,756 cases and 329,443 controls of European descent (excluding 23andMe participants) obtained from https://atlas.ctglab.nl/traitDB/4293. Depression was defined as either having a diagnosis of Major Depressive Disorder based on clinical interviews, electronic healthcare records, or self-report, and based on self-reported help-seeking or symptoms related to a broad depression phenotype.

*Anxiety disorder.* GWAS summary statistics of 25,453 anxiety cases and 58,113 controls were downloaded from https://www.kcl.ac.uk/people/kirstin-purves.[8] Participants were those who responded to the online mental health questionnaire in the UK Biobank. Cases were identified based on self-reported lifetime diagnosis of: Anxiety, nerves or generalised anxiety disorder; social anxiety or social phobia; agoraphobia; any other phobia (e.g. disabling fear of heights or spiders); and panic attacks. Additional cases were identified based on CIDI criteria for lifetime generalised anxiety disorder. Cases were excluded if they indicated a lifetime diagnosis of schizophrenia, bipolar disorder, anorexia nervosa, bulimia nervosa, any other type of psychosis or psychotic illness, autism, Asperger's or autistic spectrum disorder, and attention deficit or attention deficit and hyperactivity disorder (ADHD).

*Neuroticism*. Results from the neuroticism GWAS[9] were downloaded from https://atlas.ctglab.nl/traitDB/3795. The GWAS was performed on 390,278 European participants from the UK Biobank and the Genetics of Personality Consortium (summary statistics did not include 23andMe participants). In the UK Biobank, neuroticism was assessed using 12 items from the Eysenck Personality Questionnaire Revised Short Form (EPQ-RS)[10] and in the Genetics of Personality Consortium with 12 items from the NEO-FFI.[11]

*Intelligence.* The intelligence meta-GWAS[12] was conducted on 269,867 participants of European decent. This meta-analysis included GWAS on phenotypes relating to different domains of cognitive functioning, which were assessed using a range of standard neurocognitive tests. Summary statistics are available from https://ctg.cncr.nl/software/summary_statistics.

*Educational attainment*. Summary statistics for Educational attainment[13] was downloaded from GWASatlas available here: https://atlas.ctglab.nl/traitDB/3409. The GWAS was conducted on 318,526 UK Biobank participants and educational attainment was assessed as the highest level of education obtained.

*Alcohol use.* Summary statistics for alcohol use[14] were downloaded from https://atlas.ctglab.nl/traitDB/4069. The GWAS was conducted on 414,343 UK Biobank participants of European descent (excluding 23andMe samples). Alcohol use was coded as the average number of alcoholic drinks consumed per week.

*Smoking*. Smoking was a continuous measure coded based on a lifetime smoking index[15] devised to capture the heaviness and duration of tobacco smoking and included non-smokers ($N = $ 462,690). GWAS summary statistics[16] were obtained from https://data.bris.ac.uk/data/dataset/10i96zb8gm0j81yz0q6ztei23d.

*Cannabis use.* Summary statistics for a meta-GWAS of lifetime cannabis use[17] was downloaded from https://www.ru.nl/bsi/research/group-pages-0/substance-use-addiction-food-saf/vm-saf/genetics/international-cannabis-consortium-icc/. This GWAS was conducted on samples from the International Cannabis Consortium (ICC) and on the UK Biobank.

Summary results used in the analyses excluded 23andMe participants (43,380 cases and 118,702 controls). Cannabis use was assessed from self-reported items that asked participants to indicate whether they have ever used cannabis (yes/no).

Further details about these summary statistics can be obtained from the original GWAS publications.

***MoBa genotype data and quality control***

This study used quality controlled genotypic data from the Norwegian Mother, Father and Child cohort (MoBa), as described in full elsewhere[18]. In brief, the cohort consisted of approximately 17,000 family trios and was genotyped in three batches: 1) The NTNU Genomics Core Facility (Trondheim, Oslo) batch included 20,664 individuals and 542,585 SNPs genotyped using the Illumina HumanCoreExome (Illumina, San Diego, USA) genotyping array, version 12 1.1; 2) The second batch (12,874 individuals and 547,644 SNPs) was genotyped at the NTNU Genomics Core Facility (Trondheim, Oslo) using the Illumina HumanCoreExome (Illumina, San Diego, USA) genotyping array, version 24 1.0; 3) The ERASMUS MC (the Netherlands) batch included 17,949 individuals and 692,367 SNPs genotyped using the Illumina Global Screening Array (Illumina, San Diego, USA) version 24 1.

Quality control was performed using PLINK 1.9.[19] SNPs reported to be problematic by the Cohorts for Heart and Aging Research in Genomic Epidemiology (CHARGE) consortium and Psychiatric Genomics Consortium (PGC) were removed and duplicate samples excluded. Quality control was conducted separately in parents and offspring, and by genotyping array. Participants with a genotyping call rate less than 95% or autosomal heterozygosity greater than four standard deviations from the sample mean were removed. Ambiguous SNPs and SNPs with a genotyping call rate < 98%, minor allele frequency (MAF) < 1%, or Hardy-Weinberg equilibrium P-value $< 1 \times 10^{-6}$ were excluded. Population stratification was assessed based on principal component analysis using the HapMap3 reference panel, and non-European individuals were excluded. Individuals with a genotyping call rate < 98% or autosomal heterozygosity greater than four standard deviations from the sample mean were then removed. Biological sex was confirmed based on X chromosome heterozygosity, and any discrepancies with reported sex were flagged. Relatedness was assessed by flagging one individual from each pairwise comparison of identity-by-descent with $\hat{\pi} > 0.1$. Parent and offspring datasets were then merged into one dataset per batch, retaining the SNPs that passed quality control in both datasets. Individuals previously flagged or excluded as a duplicate, ethnic outlier, having a sex discrepancy, or high level of relatedness were retained in the merged dataset.

Quality control on the merged datasets included concordance checks on validated duplicates, excluding duplicate, tri-allelic and discordant SNPs, and again excluding individuals and SNPs with a genotyping call rate below 98%. Duplicate samples previously removed from the samples were again excluded from the merged datasets. Mendelian errors identified by the assessment of duos and trios were then recoded to missing. Insertions and deletions were also excluded.

Following the QC procedures, the Human Core Exome 12 batch included 20,231 individuals and 384,855 SNPs, the Human Core Exome 24 batch comprised 12,757 individuals and 396,189 SNPs, and the Global Screening Array batch 17,742 individuals and 568,275 SNPs. Phasing was conducted in Shapeit 2 following the duoHMM approach to account for pedigree structure. Genotype imputation was performed on the Sanger Imputation Server using the HRC release 1-1 reference panel and the Positional Burrows-Wheeler Transform (PBWT). Phasing and imputation were conducted separately for each genotyping batch. Post imputation quality control was performed by initially converting the dosages to best-guess genotypes. Individuals were removed if they had a genotyping call rate < 99% or were of non-European ethnicity. SNPs with INFO < 0.8, genotyping call rate < 98%, MAF < 1%, or a Hardy-Weinberg equilibrium P-value < $1 \times 10^{-6}$ were removed. Mendelian errors were set to missing. Relatedness, which was accounted for within generation and genotyping batch during pre-imputation QC as described above, was assessed intergenerationally and across batches by flagging one individual from each pairwise comparison of identity-by-descent with $\hat{\pi} > 0.15$ (excepting known parent-offspring relationships). Individuals were flagged for removal only if the other member of their pair would otherwise be included in the same analysis. One individual from each pair was flagged at random, except when retaining one individual in a pair would keep more duo/trio data intact than the other, in which case the other member was dropped. After quality control, a core homogeneous sample of European ethnicity (based on PCA of markers overlapping with available HapMap markers) individuals across all batches and arrays were available for use in analysis (totals prior to analysis-specific exclusions for relatedness: $N_{children} = 15,208$; $N_{mothers} = 14,804$; $N_{fathers} = 15,198$).

For the current study, we used the exclusion flags as described above to exclude one individual within pairs of relatives within and across generations and across genotyping batches, based on $\hat{\pi} > 0.15$ using a pedigree structure built in King using the following parameters: Minor allele frequency (MAF) < 1%, genotypic call rate < 98%, a maximum rate of per-individual missingness of 98% and Hardy-Weinberg equilibrium p-value < 1 x 10-6, setting all Mendelian errors to missing. Relatives were excluded at random but prioritising those with complete genetic data within mother-father-offspring trios. After exclusions, the eligible sample with available genetic data consisted of 11,262 trios (Supplementary Table 2).

**Supplementary Table 1**. Number of families with available phenotypic and genetic data.

| | Genetic data available for full trios | | |
|---|---|---|---|
| | **Yes** | **No** | **Total** |
| **Trios with child ADHD measures available** | 5405 | 2341 | 7746 |
| **Trios with missing child ADHD measures** | 5857 | 3841 | 9698 |
| **Total** | 11262 | 6182 | 17,444 |

*Note:* ADHD = Attention-deficit/hyperactivity disorder.

**Supplementary Table 2**. Correlations between trio's polygenic scores for each risk factor (*N* = 5405).

| **ADHD** | Child | Father | Mother |
|---|---|---|---|
| Child | 1.00 | 0.47 | 0.50 |
| Father | 0.47 | 1.00 | -0.02 |
| Mother | 0.50 | -0.02 | 1.00 |

| **Depression** | Child | Father | Mother |
|---|---|---|---|
| Child | 1.00 | 0.50 | 0.50 |
| Father | 0.50 | 1.00 | 0.00 |
| Mother | 0.50 | 0.00 | 1.00 |

| **Alcohol use** | Child | Father | Mother |
|---|---|---|---|
| Child | 1.00 | 0.49 | 0.51 |
| Father | 0.49 | 1.00 | 0.02 |
| Mother | 0.51 | 0.02 | 1.00 |

| **EA** | Child | Father | Mother |
|---|---|---|---|
| Child | 1.00 | 0.56 | 0.55 |
| Father | 0.56 | 1.00 | 0.09 |
| Mother | 0.55 | 0.09 | 1.00 |

| **Anxiety disorder** | Child | Father | Mother |
|---|---|---|---|
| Child | 1.00 | 0.50 | 0.48 |
| Father | 0.50 | 1.00 | -0.02 |
| Mother | 0.48 | -0.02 | 1.00 |

| **Intelligence** | Child | Father | Mother |
|---|---|---|---|
| Child | 1.00 | 0.51 | 0.52 |
| Father | 0.51 | 1.00 | 0.04 |
| Mother | 0.52 | 0.04 | 1.00 |

| **ASD** | Child | Father | Mother |
|---|---|---|---|
| Child | 1.00 | 0.49 | 0.50 |
| Father | 0.49 | 1.00 | 0.01 |
| Mother | 0.50 | 0.01 | 1.00 |

| **Neuroticism** | Child | Father | Mother |
|---|---|---|---|
| Child | 1.00 | 0.49 | 0.48 |
| Father | 0.49 | 1.00 | 0.00 |
| Mother | 0.48 | 0.00 | 1.00 |

| **Bipolar disorder** | Child | Father | Mother |
|---|---|---|---|
| Child | 1.00 | 0.49 | 0.51 |
| Father | 0.49 | 1.00 | 0.02 |
| Mother | 0.51 | 0.02 | 1.00 |

| **Schizophrenia** | Child | Father | Mother |
|---|---|---|---|
| Child | 1.00 | 0.50 | 0.51 |
| Father | 0.50 | 1.00 | 0.01 |
| Mother | 0.51 | 0.01 | 1.00 |

| **Cannabis use** | Child | Father | Mother |
|---|---|---|---|
| Child | 1.00 | 0.49 | 0.49 |
| Father | 0.49 | 1.00 | 0.01 |
| Mother | 0.49 | 0.01 | 1.00 |

| **Smoking** | Child | Father | Mother |
|---|---|---|---|
| Child | 1.00 | 0.51 | 0.52 |
| Father | 0.51 | 1.00 | 0.07 |
| Mother | 0.52 | 0.07 | 1.00 |

*Note:* ADHD = Attention-deficit/hyperactivity disorder; ASD = Autism spectrum disorder; EA = Educational attainment.

**Supplementary Table 3**. Comparison of mean standardized polygenic scores between families with and without child ADHD scores available.

| PGS | Role | Mean polygenic scores: Child phenotype available | Mean polygenic scores: Child phenotype missing | t | df | p |
|---|---|---|---|---|---|---|
| ADHD | Child | -0.0263 | 0.0246 | 3.09 | 14722 | 0.002 |
| | Father | -0.0173 | 0.0142 | 1.89 | 14544 | 0.058 |
| | Mother | -0.0396 | 0.0329 | 4.31 | 14197 | $1.61 \times 10^{-5}$ |
| Autism spectrum disorder | Child | 0.0102 | -0.0095 | -1.20 | 14722 | 0.231 |
| | Father | 0.0129 | -0.0105 | -1.41 | 14544 | 0.159 |
| | Mother | 0.0082 | -0.0068 | -0.89 | 14197 | 0.373 |
| Schizophrenia | Child | -0.0321 | 0.0300 | 3.78 | 14722 | $1.56 \times 10^{-4}$ |
| | Father | -0.0258 | 0.0211 | 2.83 | 14544 | 0.005 |
| | Mother | -0.0537 | 0.0445 | 5.85 | 14197 | $5.17 \times 10^{-9}$ |
| Bipolar disorder | Child | -0.0090 | 0.0080 | 1.06 | 14722 | 0.289 |
| | Father | -0.0058 | 0.0048 | 0.64 | 14544 | 0.523 |
| | Mother | -0.0207 | 0.0172 | 2.26 | 14197 | 0.024 |
| Depression | Child | -0.0142 | 0.0133 | 1.66 | 14722 | 0.096 |
| | Father | -0.0192 | 0.0157 | 2.10 | 14544 | 0.036 |
| | Mother | -0.0258 | 0.0214 | 2.81 | 14197 | 0.005 |
| Anxiety | Child | -0.0099 | 0.0093 | 1.17 | 14722 | 0.243 |
| | Father | -0.0082 | 0.0067 | 0.90 | 14544 | 0.370 |
| | Mother | -0.0020 | 0.0017 | 0.22 | 14197 | 0.825 |
| Neuroticism | Child | -0.0212 | 0.0198 | 2.49 | 14722 | 0.013 |
| | Father | -0.0092 | 0.0075 | 1.01 | 14544 | 0.314 |
| | Mother | -0.0123 | 0.0102 | 1.33 | 14197 | 0.183 |
| Intelligence | Child | 0.0455 | -0.0425 | -5.35 | 14722 | $9.12 \times 10^{-8}$ |
| | Father | 0.0393 | -0.0321 | -4.31 | 14544 | $1.66 \times 10^{-5}$ |
| | Mother | 0.0539 | -0.0447 | -5.87 | 14197 | $4.40 \times 10^{-9}$ |
| Educational attainment | Child | 0.0295 | -0.0276 | -3.52 | 14722 | $4.32 \times 10^{-4}$ |
| | Father | 0.0208 | -0.0170 | -2.32 | 14544 | 0.020 |
| | Mother | 0.0357 | -0.0296 | -4.00 | 14197 | $6.26 \times 10^{-5}$ |
| Alcohol use | Child | -0.0096 | 0.0097 | 1.13 | 14722 | 0.260 |
| | Father | 0.0049 | -0.0040 | -0.53 | 14544 | 0.594 |
| | Mother | -0.0062 | 0.0051 | 0.67 | 14197 | 0.502 |
| Smoking | Child | -0.0531 | 0.0496 | 6.26 | 14722 | $4.02 \times 10^{-10}$ |
| | Father | -0.0390 | 0.0319 | 4.27 | 14544 | $1.94 \times 10^{-5}$ |
| | Mother | -0.0604 | 0.0501 | 6.59 | 14197 | $4.52 \times 10^{-11}$ |
| Cannabis use | Child | 0.0109 | -0.0102 | -1.29 | 14722 | 0.199 |
| | Father | 0.0110 | -0.0090 | -1.20 | 14544 | 0.230 |
| | Mother | 0.0063 | -0.0052 | -0.69 | 14197 | 0.493 |

*Note:* ADHD = Attention-deficit/hyperactivity disorder.
